# Beyond prescription: Determinants of low-dose aspirin adoption and uptake across the pre-eclampsia prevention pathway in Nepal – a qualitative study guided by the COM-B model

**DOI:** 10.64898/2026.11.13.26368074

**Authors:** Leila Harrison, Sushmita Uprety, Anusha Ojha, Trishna Rai, Pratiksha Paudel, Jaya Dhungana, Suchitra Chandrasekaran, Usha Ramakrishnan, Archana Shrestha, Elizabeth C. Rhodes

## Abstract

Despite strong evidence supporting low-dose aspirin (LDA) for preventing pre-eclampsia among women at risk, implementation remains suboptimal globally. The prevention pathway encompasses early antenatal care engagement, screening and risk identification, prescribing LDA, and medication uptake. This qualitative study explored barriers and facilitators to LDA adoption by providers (i.e., prescribing) and uptake among women in Nepal.

A qualitative study was conducted with pregnant women, healthcare providers, and maternal and child health stakeholders in Nepal. Data collection and analysis were guided by the Capability, Opportunity, Motivation, and Behavior (COM-B) model. Participants were purposively recruited from three tertiary hospitals and maternal health organizations. Semi-structured interviews explored experiences and perspectives related to antenatal care, such as pre-eclampsia risk screening, LDA prescribing, and uptake. Data were analyzed using rapid qualitative analysis and organized according to the COM-B components.

Seventy-three participants were interviewed, including 36 pregnant women, both at risk and not at risk for pre-eclampsia; 29 healthcare providers, including consultant/senior obstetricians, mid-level doctors, and nurses or midwives; and 8 maternal and child health stakeholders representing government, multi-lateral, non-governmental, professional, and academic institutions. LDA adoption and uptake were affected by interdependent gaps across the prevention pathway. Delayed antenatal care engagement and inconsistent screening limited timely identification of women at risk, while gaps in national guidance and standardized prescribing protocols contributed to variable provider prescribing practices. Among women prescribed LDA, clear counselling on its purpose and safety and trust in providers supported uptake.

LDA adoption and uptake depend on a functioning prevention pathway in which women engage with antenatal care early enough for timely risk assessment, those at risk are identified through screening, providers prescribe appropriately, and women are supported to use the medication. Strengthening this pathway requires coordinated attention to health-system and provider capacity as well as communication with women, rather than strategies focused narrowly on prescribing or medication use. These findings will inform the development of implementation strategies for Nepal and other low-resource settings seeking to integrate evidence-based preventive interventions into maternal healthcare systems.

## Introduction

Hypertensive disorders during pregnancy (HDP) are a leading cause of maternal and fetal morbidity and mortality worldwide, with the greatest burden borne by low-and middle-income countries (LMICs) [1–2]. Globally, HDP affect an estimated 10-22% of pregnant women, with prevalence rising over recent decades alongside increases in maternal age and the prevalence of pre-existing medical conditions [3]. Pre-eclampsia, one of the most severe forms of HDP, is responsible for approximately 46,000 maternal deaths and 500,000 fetal or newborn deaths annually and contributes substantially to adverse birth outcomes, including preterm birth, fetal growth restriction, stillbirth, and neonatal mortality [4,5]. In LMICs, where access to timely diagnosis and high-quality obstetric care may be limited, the consequences of pre-eclampsia are particularly severe. In Nepal, pre-eclampsia accounts for 22% of maternal deaths, and is associated with 25% of preterm births and 38% of low-birth-weight infants [6–8]. Beyond its immediate effects on pregnancy outcomes, pre-eclampsia has important long-term implications for both mother and child. Women who develop pre-eclampsia face elevated lifetime risks of hypertension, kidney disease, metabolic disorders, cardiovascular disease and dementia, while infants born preterm or growth-restricted due to pre-eclampsia are at increased risk of non-communicable diseases later in life [9,10]. Given this substantial burden, identifying women at risk and the use of effective preventive interventions are critical, particularly in LMICs where limited capacity for disease management, combined with already constrained health systems, underscores the importance of prevention.

Extensive evidence demonstrates that low-dose aspirin (LDA) reduces the risk of pre-eclampsia among women at moderate- to high-risk, particularly when initiated by 12 weeks’ gestation [11–16]. In addition to its clinical benefits, LDA has been shown to significantly reduce healthcare costs related to pre-eclampsia [17–19]. Consequently, the World Health Organization (WHO) and numerous international guideline bodies recommend LDA for pre-eclampsia prevention in at-risk pregnancies [15,20].

First-trimester screening for pre-eclampsia risk during antenatal care (ANC) provides an important opportunity to identify women who may benefit from preventive interventions, including LDA [21,22]. While women may attend ANC and complete the recommended number of visits [23], delayed initiation of ANC after 12 weeks’ gestation can prevent timely risk identification and initiation of LDA within the effective gestational window. Thus, despite strong evidence and global recommendations, the effectiveness of LDA depends on successful implementation across a prevention pathway comprising four behaviors: women’s early engagement with ANC, healthcare provider screening and risk assessment, healthcare provider adoption of LDA, and women’s uptake of LDA. Provider adoption refers to incorporating appropriate LDA prescribing into clinical practice for women at risk, while uptake refers to women initiating and using LDA. Breakdowns at any stage can limit its impact, with suboptimal implementation widely documented globally, particularly in LMICs [24–32].

In Nepal, LDA is included on the WHO’s list of essential medicines [33] and is widely available [34], but it has not yet been incorporated into the National Safe Motherhood Program guidelines [35,36] – the country’s leading initiative to reduce maternal and fetal morbidity and mortality. As evidence supporting LDA has grown, representatives of the Nepal Ministry of Health and Food Safety (MOHFS) have expressed a need for evidence on the most effective strategies to support its adoption and uptake (personal communications, 2024-2026).

To address this gap, the objective of this study was to identify barriers and facilitators to LDA adoption and uptake across the pre-eclampsia prevention pathway in Nepal, from the perspectives of pregnant women, healthcare providers, and key maternal and child health (MCH) stakeholders. The findings provide context-specific insights that will inform the co-design of implementation strategies and lay the foundation for future implementation efforts in Nepal.

## Materials and Methods

### Theoretical Framework

This study was guided by the Capability, Opportunity, Motivation, and Behavior (COM-B) model, which proposes that behavior is dependent on capability, opportunity, and sufficiently strong motivation [37]. Capability refers to the physical or psychological capacity to perform a behavior; opportunity to the physical or social factors that enable or constrain behavior; and motivation to reflective or automatic processes that influence the behavior [37].

COM-B was selected for its simplicity and comprehensiveness, enabling consistent analysis of barriers and facilitators to LDA adoption and uptake across multiple actors and stages of the prevention pathway. Although originally developed to understand individual behavior, its conceptual parsimony allows application across multiple health system actors, making it well suited to examining complex implementation processes such as ANC delivery in low-resource settings. The framework informed both the development of interview guides and the analysis, organization, and presentation of findings.

### Study team and reflexivity statement

This study was conducted through a partnership between researchers at Emory University (USA) and Kathmandu University School of Medical Sciences (Nepal) with expertise in maternal health, implementation science, and qualitative methods. Guided by a critical realist perspective, the study assumes that real-world mechanisms influence behavior, while participants’ accounts provide partial, socially situated insights [38]. The research team included Nepali and non-Nepali researchers, with several team members bilingual in Nepali and English. Participants were informed of the researchers’ institutional affiliations and roles in the study.

Given LH’s position as an external researcher, the team remained attentive to potential power differentials and the possibility that providers could perceive the research as evaluative. Reflexivity was supported through open-ended, neutral questioning and emphasis on understanding participants’ experiences, as well as ongoing team discussions to examine positionality, power dynamics, and how researchers’ assumptions and prior knowledge might shape emerging interpretations [39]. Interpretation of findings involved LH and Nepal-based team members (SU, AO, TR, PP), incorporating both non-Nepali and Nepali perspectives and contextual knowledge.

### Study design, sampling, and recruitment

This study was reported in accordance with the Consolidated Criteria for Reporting Qualitative Research (COREQ) [40]. Purposive sampling was used to recruit pregnant women and healthcare providers from three tertiary hospitals in Nepal that were selected for high patient volume of pregnant women, advanced obstetric care, and variety in hospital type (e.g., community-based, government-run, and private).

Pregnant women ≥ 18 years of age and ≥ 20 weeks’ gestation were recruited during ANC visits while awaiting their appointments. A screening form was used to assess eligibility, including age, gestational age, and pre-eclampsia risk status. Eligible women included both those with pre-eclampsia risk factors (as defined by WHO criteria [20]) and uncomplicated pregnancies to capture a range of experiences. Risk status was determined through self-report and ANC card review, with ANC cards prioritized in cases of discrepancy. Healthcare providers, including obstetricians, residents, nurses, and midwives involved in ANC and high-risk pregnancy management, were recruited via departmental heads or senior clinical focal persons, with support from nursing and administrative staff to identify eligible participants across professional roles and levels of clinical experience.

To capture broader system-level perspectives, key stakeholders involved in MCH policy and program implementation in Nepal were included. Participants were identified through professional networks and snowball sampling, beginning with established contacts within the Government of Nepal MOHFS and expanding to include representatives from multilateral organizations and non-governmental implementing partners.

Recruitment timelines differed across participant groups according to interviewer availability. A single researcher conducted all interviews with pregnant women, and recruitment took place sequentially across the three participating hospitals: Dhulikhel Hospital (12/11/2024-11/12/2024), Paropakar Maternity Hospital (23/01/2025-15/07/2025), and Kathmandu Medical College (01/08/2025-03/09/2025). In contrast, healthcare provider recruitment occurred concurrently across all three hospitals from October to November 2025, with interviews conducted by two researchers. MCH stakeholders were recruited between 17/11/2025 and 25/03/2026.

### Data collection and analysis

Written informed consent was obtained from all participants prior to data collection. Semi-structured interview guides were developed for each participant group in English, translated into Nepali by a native Nepali-speaking team member, and back translated to ensure conceptual equivalence and preserve meaning across languages. Guides covered ANC experiences, pre-eclampsia risk screening, and prevention, including LDA, and were piloted with three participants and iteratively refined prior to formal data collection; pilot interviews were not included in the analysis.

In-depth interviews with pregnant women were conducted in private hospital spaces and lasted approximately 60 minutes. Key informant interviews with healthcare providers were also conducted in private hospital spaces and lasted approximately 30 minutes. Key informant interviews with MCH stakeholders were conducted either in person or virtually via Zoom, depending on participant preference and feasibility, and lasted approximately 45 minutes. Interviews were conducted in English or Nepali according to participant language preference. SU conducted all interviews with pregnant women in Nepali; LH conducted interviews with doctors and MCH stakeholders in English, while AO conducted interviews with nurses and midwives in Nepali. LH provided training in best practices for recruitment and qualitative data collection and analysis, including interviewing, probing, rapid qualitative analysis (RQA), and COM-B-guided data summarization and mapping, with ongoing supervision and support throughout the research process [39].

All interviews were audio recorded. Interviews were transcribed verbatim in the language in which they were conducted by Nepali team members and external contractors. Transcripts of interviews conducted in Nepali were subsequently translated into English for analysis. Weekly debrief sessions throughout data collection were used to practice reflexivity by providing opportunities for team members to reflect on interpretations and challenge assumptions regarding ANC experiences and pre-eclampsia prevention. These sessions also informed iterative refinement of the interview guides including the addition of probes to delve deeper into topics that arose in previous interviews, and identification of emergent themes [39,41]. Concurrent analysis supported an iterative assessment of whether sufficient data had been collected to address the study objectives [41–45]. This assessment considered whether additional interviews were generating new substantive themes, indicating thematic saturation, as well as the breadth of perspectives represented across participant groups and hospital sites. Recruitment continued iteratively until no new substantive themes emerged and the research team judged that sufficient data had been collected to address the study objectives [41–45].

Data analysis followed a framework-guided RQA approach [46], to facilitate systematic and transparent data reduction and synthesis within a multidisciplinary team and to support iterative analysis during data collection. Analysis was led by LH, with input from ECR and AS, and conducted with Nepali team members (SU, AO, PP, and TR), who supported transcript summarization and mapping. Weekly team discussions supported consistency and resolution of differences in interpretation, with LH reviewing and finalizing the summaries, mapping, and narrative synthesis. Structured Excel templates were used to summarize transcripts by interview question, with deductive coding guided by COM-B. Analysts also applied inductive coding to capture emergent concepts not predefined by the framework, comparing these against COM-B components to assess fit. Illustrative quotations were extracted alongside summaries to preserve linkage to the raw data. The template was piloted with five transcripts and refined iteratively prior to use with the full dataset.

Next, barriers and facilitators were mapped by participant group and COM-B component from the completed summaries into an Excel matrix followed by narrative synthesis. All identified determinants could be meaningfully mapped to a COM-B component. Findings were organized along the pre-eclampsia prevention pathway, conceptualized as a sequential and interdependent process comprised of four behaviors: 1) women’s early engagement with ANC, 2) healthcare provider screening and risk assessment, 3) healthcare provider adoption (i.e., prescribing) of LDA, to 4) women’s uptake of LDA. Within each stage, determinants (i.e., barriers and facilitators) were analyzed and organized by COM-B components across patient-, provider-, and health-system levels.

### Ethical considerations

Written informed consent was obtained from all participants prior to data collection. Ethical approval for this study was obtained from the Ethical Review Board of the Nepal Health Research Council (No. 676/2023) and the Emory University Institutional Review Board (No. 2025P013433). The study was conducted in accordance with the principles of the Declaration of Helsinki.

## Results

We interviewed 73 participants, including 36 pregnant women, 29 healthcare providers, and 8 MCH stakeholders (Table 1). All pregnant women reported being literate, and the largest proportion had a bachelor’s or post-graduate degree (41.6%); 44.4% reported having any form of health insurance. Among eligible women who had risk factors for pre-eclampsia, only approximately half (52.6%) self-reported or had documentation of LDA use in their ANC cards. Healthcare providers included consultant or senior obstetric doctors, mid-level doctors, and nurses or midwives. Nearly two-thirds were doctors, and only they had prescribing authority. MCH stakeholders represented government, multilateral, non-governmental, professional, and academic institutions.

**Table 1.** Characteristics of participants interviewed (N=73)

| **Characteristics** | **Value** |
|---|---|
| ***Pregnant women (n=36)*** |  |
| Age (years) | 29.9 (SD 4.6) |
| Gestational age (weeks) | 29.6 (SD 5.7) |
| Parity (≥1 birth) | 17 (47.2%) |
| Education level |  |
| No formal schooling | 1 (2.8%) |
| Primary or secondary school | 10 (27.8%) |
| High school | 10 (27.8%) |
| Bachelor’s or post-graduate | 15 (41.6%) |
| Literacy |  |
| Can read | 36 (100%) |
| Can write | 36 (100%) |
| Any form of health insurance | 16 (44.4%) |
| Pregnancy risk status |  |
| At risk | 19 (52.8%) |
| Taking LDA | 10 (52.6%) |
| Not taking LDA | 9 (47.4%) |
| Not at risk | 17 (47.2%) |
| ***Healthcare providers (n=29)*** |  |
| Professional role |  |
| Consultant/Senior obstetrician | 11 (37.9%) |
| Mid-level doctors (residents/registrars/medical officers/SHO) | 7 (24.2%) |
| Nurses and midwives | 11 (37.9%) |
| Years of clinical experience in MCH |  |
| <10 years | 14 (48.3%) |
| 10-19 years | 11 (37.9%) |
| ≥20 years | 4 (13.8%) |
| Prescribing capability |  |
| Yes | 18 (62.1%) |
| No | 11 (37.9%) |
| ***MCH Stakeholders (n=8)*** |  |
| Organizational affiliation |  |
| Government/Ministry of Health | 2 (25.0%) |
| Professional medical society | 1 (12.5%) |
| Multilateral organizations | 2 (25.0%) |
| Non-governmental organizations | 2 (25.0%) |
| Academic institution | 1 (12.5%) |
| Years of professional MCH experience |  |
| <10 years | 2 (25.0%) |
| 10-19 years | 4 (50.0%) |
| ≥20 years | 2 (25.0%) |

*Footnotes:* Values are presented as mean (standard deviation) for continuous variables and n (%) for categorical variables. SHO, Senior Health Officer; MCH, Maternal and Child Health.

Findings are presented by key behaviors in the pre-eclampsia prevention pathway (Fig 1). The results focus on key determinants, selected based on their recurrence across interviews and participants’ emphasis on their role in explaining the target behaviors, while the complete set of identified determinants is presented in S1 Tables 1-4.

**Fig 1.**
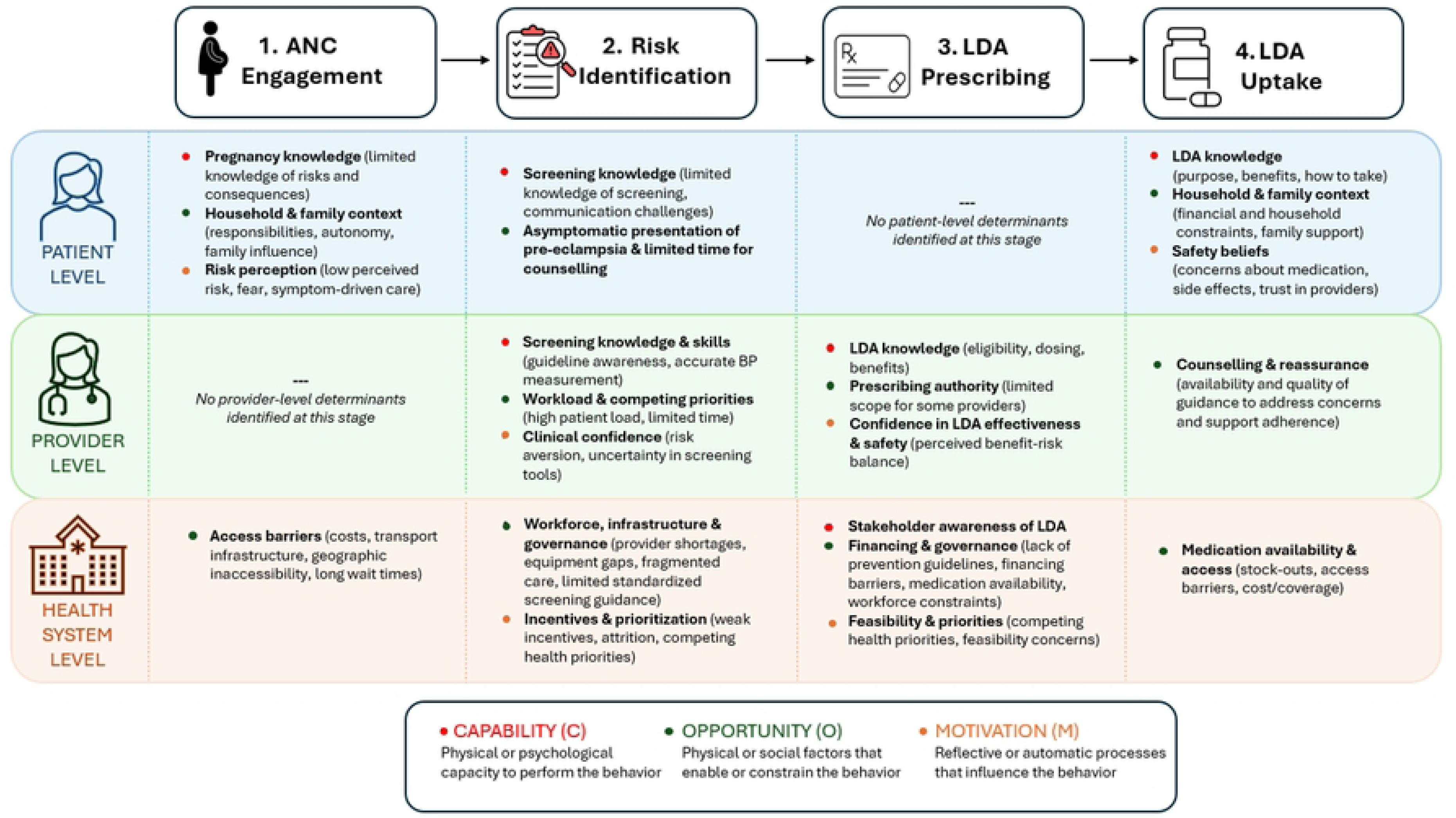
Conceptual diagram of key determinants across the pre-eclampsia prevention pathway by level and COM-B component.

### Determinants of ANC engagement

Early engagement with ANC was shaped by interacting *patient-* and *health-system level* determinants across COM-B components.

From a **psychological capability** perspective at the *patient-level,* limited knowledge of pregnancy risks, particularly pre-eclampsia, its symptoms, and consequences, combined with low health literacy reduced women’s ability to recognize the need for early ANC. Many reported having no knowledge of pre-eclampsia, stating: *“I don’t know anything about it.”*

Within **physical opportunity**, all participants described some combination of both *health-system level* structural barriers that contributed to delayed or inconsistent ANC attendance, including costs of care and transport, geographic inaccessibility, and limited transport infrastructure, and *patient-level* barriers, such as time constraints related to household responsibilities and seasonal work and concerns about difficult or unsafe travel conditions. As one woman explained:

*“In our village, there are not many transportation facilities. It takes at least an hour to get here. In addition, I have a small child at home, so I would have to leave them behind to come here. That is what makes it inconvenient”* [Pregnant woman #13].

Conversely, participants described reduced costs of care (e.g., existing financial assistance mechanisms) as facilitating ANC engagement.

At the *patient-level* within **social opportunity**, gender norms and household power dynamics rooted in broader gender inequalities limited women’s autonomy in care-seeking. While pregnancy care was often viewed as solely a woman’s responsibility, decision-making and financial control were frequently held by male partners or senior family members. For example, one MCH stakeholder shared:

> *“And they [women] may take some time to come…There are so many barriers, we call it you know delays…The first delays would be inside the household, because the woman, because of the nature of the society, patriarchal society, the women do not decide on themselves”* [MCH stakeholder #6].

Limited partner and family support, along with cultural beliefs and stigma (e.g., surrounding adolescent pregnancy), were described as factors that could further delay ANC initiation by making it difficult for women to attend without assistance and, for some, creating concerns about disclosing their pregnancy and facing judgment or discrimination.

At the *patient-level* within **reflective motivation**, many women reported low perceived risk and seriousness of pregnancy, which reduced the perceived need for ANC and information on conditions such as pre-eclampsia. This was often linked to absence of personally experiencing symptoms or prior experience, leading to a more reactive approach to health in which information was only sought if problems arose. Conversely, prior experiences with high blood pressure during pregnancy increased perceived risk and facilitated ANC engagement. Within **automatic motivation**, emotional responses to pregnancy risks, including fear, also shaped engagement with information and care. Concern about complications increased vigilance for some women. One woman described becoming very frightened after learning about pre-eclampsia online, stating, “*these days it feels like I don’t even want to hear such news, just hearing about it makes me very scared. It has kind of stayed in my mind.*” This distress prompted her to want her blood pressure checked. For other women, however, frightening information discouraged care-seeking, as concerns about potential complications or negative outcomes created stress and contributed to avoidance of ANC.

Participants proposed additional facilitators spanning **psychological capability** and **social opportunity**, including greater knowledge of pregnancy risks and the importance of early and continued ANC, and family, community, and peer support that normalized ANC and increased awareness of pre-eclampsia.

Delayed engagement was also noted to increase reliance on higher-cost screening and diagnostic care later in pregnancy, in addition to limiting the opportunity to initiate preventive strategies such as LDA within the effective gestational window.

### Determinants of provider screening and risk identification during ANC encounters

Following ANC engagement, screening and risk identification during ANC encounters represented a critical but often incomplete step in the prevention pathway, as entry into care alone did not ensure that women eligible for LDA were identified within the relevant gestational window. Thus, screening and risk identification were inconsistent across and within facilities, reflecting the combined influence of *patient-, provider-,* and *health-system level* determinants across COM-B components.

**Psychological capability** determinants were evident at both *provider-* and *patient-levels*. At the *provider-level*, knowledge of pre-eclampsia, including risk factors, consequences, screening approaches, and international guidelines was generally strong. However, most providers lacked awareness of Nepal’s *National Medical Standard for Maternal and Newborn Care (2022),* which includes recommendations for blood pressure measurement and urine testing to identify pre-eclampsia risk. In addition, limited knowledge of correct blood pressure measurement techniques among frontline nurses, who are primarily responsible for routine measurement, reduced screening reliability, with inaccurate results often requiring re-check by doctors:

> *“Sometimes the nursing student write whatever blood pressure. So if we think that she [pregnant woman] is having some risk then we’ll take blood pressure ourselves. And sometimes the nurse blood pressure is something and our blood pressure is different”* [Doctor #6].

At the *patient-level*, understanding of pre-eclampsia and screening was constrained by communication barriers, including limited shared language, use of technical terminology, and inconsistent counselling. For example, several healthcare providers noted that the absence of a clear Nepali term for pre-eclampsia made it difficult to explain the condition to patients:

> *“There is no proper word for pre-eclampsia. We have to explain in English because we haven’t that proper term…so when I say that you might have pre-eclampsia, then they don’t understand what is pre-eclampsia. Then we have to explain again and they are so much confused, and they just walk away”* [Doctor #4].

Participants described information overload and exposure to conflicting externally sourced health information (e.g., from Google or ChatGPT) as determinants that could further reduce patients’ ability to interpret and act on clinical guidance.

**Physical opportunity** was a key component across *patient- and health-system levels*. At the *patient-level*, asymptomatic presentation of pre-eclampsia, brief consultations, and limited opportunities for questions constrained opportunities for early detection. Participants identified facilitators including access to educational materials about pregnancy risks, adequate time for counselling and patient-family education, and opportunities for women to ask questions and communicate openly with providers.

At the *health-system level,* physical opportunity was shaped by workforce and service delivery constraints. Shortages and uneven distribution of skilled healthcare providers, staff transfers, and rural staffing gaps limited opportunities to screen, while high patient volumes, competing clinical responsibilities, and limited protected time reduced healthcare provider’s opportunity to deliver consistent screening and counselling. As one healthcare provider described:

> *“The doctors are burned out here. Because for maybe 50, 60 patient there are only one doctor. And so we are unable to take care of them, and we are unable to give them more time…maybe we need to just for one day there need be certain number of patient. And the hospital should like to say cut off [patients]. Yeah, after certain so that not only for doctors there it’s benefit for the patient also, no? They’ll find more time to talk with the doctor”* [Doctor #6].

Participants identified facilitators including decentralized service delivery infrastructure that could enable community-based care and existing continuing medical education programs that could facilitate ongoing provider training. These *health-system level* workforce and service delivery determinants also shaped healthcare providers’ ability to prescribe LDA for at-risk women.

Participants described how screening was further constrained by equipment and infrastructure limitations, including faulty blood pressure devices, limited availability of uterine artery Doppler machines, supply chain challenges, high diagnostic costs (for both health facility procurement and patients), and inadequate space for screening services. Fragmented service delivery, particularly in peripheral facilities, disrupted continuity between screening, risk identification, and follow-up. Although uterine artery Doppler machines were available at all three facilities, access was not always consistent due to factors such as limited appointment availability within radiology services, which often delayed screening scans to subsequent visits, which many women do not return for. One healthcare provider described this challenge:

> *“Sometimes what happened is they are coming from far, so we advise them to do the scan, but the scan is already full. So they cannot get an appointment in that particular period of time…that window period of 11 to 13 weeks is gone…So, if they are coming like eight weeks, nine weeks, then okay, they can take an appointment after two weeks and do the scan. But even in sometimes like 11 to 13 weeks they are coming, we do not have the appointment you know. Already it’s already full so they’re unable to do on that particular time”* [Doctor #5].

Conversely, participants identified existing ANC platforms, including routine blood pressure measurement, urine testing for albumin, history taking, and the use of ANC cards (paper records used to document maternal health information), as well as nationally implemented programs such as *Skilled Birth Attendant training* and government incentive programs (e.g., *Aama Surakshya Programme*), as opportunities that could be leveraged to support systematic screening.

Importantly, the absence of national prevention-specific clinical guidance for pre-eclampsia and standardized hospital screening protocols were key barriers contributing to variability in screening and risk identification across and within healthcare facilities.

**Reflective motivation** further shaped screening practices across all levels, with the strongest influence at the *provider-* and *health-system levels*. At the *provider-level*, perceived blame and accountability for adverse outcomes following delayed care-seeking contributed to risk-averse practices, including earlier referral that could delay screening, while limited policy support and uncertainty regarding the diagnostic accuracy of uterine artery Doppler reduced confidence in screening approaches.

At the *health-system level,* low salaries, contract-based employment, and limited career opportunities, reduced healthcare provider motivation to engage in high-quality care and contributed to provider attrition from the public health system through migration abroad and transition to private-sector employment for better opportunities. One MCH stakeholder described how decentralized decision-making at the local level could result in government funding for nursing and midwifery positions being divided among multiple staff, reducing individual salaries and undermining motivation: “*How they can be motivated?”*

MCH stakeholders and providers described improved remuneration and in-country career opportunities as facilitators to strengthen provider motivation, thereby improving retention and workforce stability.

In addition, participants highlighted that competing health priorities within the government could potentially limit attention and resources available for establishing systematic pre-eclampsia screening. However, pre-eclampsia was widely recognized by MCH stakeholders and healthcare providers as a major contributor to maternal morbidity and mortality and was viewed as a priority area for investment, including strengthening screening and LDA integration. Despite this recognition, concerns regarding the reliability of nurse-conducted obstetric scans limited confidence in task-shifting and expanding diagnostic roles within the health system.

### Determinants of provider prescribing of LDA for at-risk pregnant women

Multiple determinants shaped prescribing behavior for LDA across *provider-* and *health-system levels* of multiple COM-B components.

From a **psychological capability** perspective, knowledge gaps and uncertainty regarding pre-eclampsia prevention and LDA use contributed to inconsistent prescribing among both *provider-* and *health-system levels*. At the *provider-level*, knowledge of LDA by doctors, including timing, dosing, safety, and eligibility criteria, functioned as both a facilitator and barrier: strong understanding supported appropriate prescribing, whereas uncertainty limited its routine and consistent use. For example, one doctor described that LDA prescription was “*not practiced here*” except by “*few doctors*” and that prescribing decisions were “*individual to them*,” noting that some providers initiated LDA among high-risk patients, including those with advanced maternal age or previous pre-eclampsia.

At the *health-system level*, similar gaps were reflected in limited familiarity with LDA as a preventive intervention among MCH stakeholders, as well as uncertainty regarding its clinical use, including timing, risks, and prescribing authority. For example, one MCH stakeholder acknowledged limited familiarity with the evidence supporting LDA effectiveness: *“Hmm I think there are evidence that it helps…but I don’t have much more than that.”*

Within **physical opportunity** at the *health-system level*, similar to screening, participants described limitations in diagnostic capacity and availability of specialized, costly tests (e.g., PLGF, β-hCG, coagulation profiles) that reduced confidence in risk assessment and safe clinical decision-making to prescribe LDA. Participants also described workforce shortages and uneven distribution of skilled providers with prescribing capacity as further limiting opportunities for LDA prescribing. Existing national guidelines support pre-eclampsia management (e.g., use of magnesium sulfate), but as noted previously, the absence of national prevention-specific guidance for pre-eclampsia also emerged as a major barrier to LDA prescribing, contributing to uncertainty around how and when LDA should be used after women are identified as being at risk. Although LDA is prescribed by some healthcare providers in tertiary care settings, it is not included in national essential medicine lists or routine government protocols, restricting standardized provision within the public health system. As one MCH stakeholder noted:

> *“In major document there is no, any provision of any recommendations for aspirin use in pregnancy. But we have heard that some hospitals, they are practicing by providing the low dose of aspirin in pregnant women…but the distal document for the country is, there is no information”* [MCH stakeholder #1].

MCH stakeholders emphasized that adoption of LDA into routine care will require alignment across existing national systems, with gaps in policy, financing, and implementation infrastructure acting as barriers to consistent prescribing. Participants identified several national systems relevant to this alignment, including ANC guidelines, the Essential Medicines List (which guides procurement and availability of medications), the Basic Health Service Package (a list of essential medicines provided free of cost), insurance schemes, and government training platforms (e.g., Skilled Birth Attendant programs and Emergency Obstetric and Newborn Care job aids). Stakeholders described such alignment as important for supporting procurement and financing, clarifying eligibility criteria and provider roles, and establishing referral pathways linking risk identification to prescribing. Inclusion within systems such as the Basic Health Service Package was viewed as particularly important because it would formally mandate government provision; however, MCH stakeholders noted that adoption would require sufficient financing not only for medication availability but also for training health workers. For example, one MCH stakeholder explained, “*It’s not only about making [LDA] available, it’s also about giving the training to all the lower health staffs…so that would be a quite budgetary aspect*.”

Stakeholders further emphasized the importance of sustainable financing mechanisms, recognizing that “*the government can only afford so much to be free,*” and suggesting that integration with social health insurance schemes could help reduce financial barriers and support long-term implementation.

In the absence of national protocols and standardized delivery mechanisms, Nepal’s decentralized health system may further contribute to variability in LDA adoption, as local differences in resources, oversight, and service delivery capacity influence how preventive interventions are operationalized. Restrictions on prescribing authority among lower-level providers (e.g., nurses and auxiliary nurse midwives) also limit decentralized delivery. One MCH stakeholder highlighted how guideline endorsement could enable broader provider roles, noting:

> *“So, right now, aspirin might be difficult because it’s not a part of a formal guideline, but then if it becomes a part of the national guideline, then it’s like there might be avenues which it might open those avenues for the nurses and midwives as well”* [MCH stakeholder #2].

**Reflective motivation** shaped LDA prescribing behavior at both *provider*- and *health-system levels*. At the *provider-level*, perceptions of LDA safety and effectiveness influenced prescribing confidence, with providers who viewed LDA as evidence-based and beneficial reporting greater willingness to prescribe. In private-sector settings, some providers perceived national guidance as less relevant to their clinical contexts, particularly when they viewed their care practices as exceeding those in the public sector, potentially contributing to variation in prescribing practices. At the *health-system level*, stakeholder willingness to integrate LDA into routine care was shaped by perceptions of evidence strength, safety, and feasibility within existing systems. A major concern noted by many participants was the capacity of peripheral facilities to manage potential complications associated with LDA use. As one MCH stakeholder noted:

> *“Developing this low-dose aspirin guideline would be feasible, but also I’m not sure to what extent it increases the possibility or incidence of postpartum hemorrhage…especially given the ability of the peripheral level health facilities and the hospitals to deal with postpartum hemorrhage”* [MCH stakeholder #2].

In addition, perceptions of feasibility were shaped by competing ANC priorities (e.g., achieving recommended ANC visit schedules) and concerns that adherence to an additional medication may be limited, given existing challenges in adherence with established free supplements such as iron-folic acid.

### Determinants of LDA uptake by pregnant women

Uptake reflects the translation of prescribing into sustained medication use and is shaped primarily by *patient-level* determinants across COM-B components, with additional influences from *provider-* and *health-system level* physical opportunity factors.

Within **psychological capability** at the *patient-level*, building on the limited knowledge of pre-eclampsia described above, women also had limited knowledge of LDA, including those at-risk for pre-eclampsia who were eligible for, and even actively taking, the medication. Many reported little or no understanding of its purpose, benefits, safety, or dosing, with some confusing LDA with other pregnancy supplements such as calcium or iron and questioning the need for additional medication. For example, one woman reported that “*there isn’t much*” that can be done to prevent high blood pressure during pregnancy and was unaware of any preventive medications. Only after the interviewer mentioned aspirin as a preventive strategy did she realize she was already taking it, explaining, “*The doctor didn’t explain the reason. When I asked, they didn’t explain. They just said to take it.*” Limited recall of clinical instructions, as well as pregnancy-related “brain fog” and forgetfulness, further constrained sustained uptake. In contrast, **physical capability** was not perceived as a barrier, as women taking LDA reported that the medication was easy to take, with no difficulty or discomfort swallowing the pill.

**Physical opportunity** influenced LDA uptake across *patient-, provider-* and *health-system levels*. At the *patient-level,* financial constraints (e.g., prioritizing food over medication), difficult travel conditions, and time constraints related to competing household responsibilities (e.g., childcare), limited women’s ability to access and sustain LDA use. Many women described experiencing multiple barriers simultaneously: “*it could be financial difficulties, bad road conditions, or longer distance from home to hospital…or a lack of someone at home to help when needed.*” At the *provider-level*, clear, simple, and structured counselling on LDA timing, dosage, purpose, benefits, and safety supported correct and sustained use, whereas limited counselling constrained uptake. One woman described receiving LDA from a provider but returning it because she did not understand why it had been prescribed or how it related to her pregnancy: “*The doctor had given it to me…I even brought it home. Then I returned that medicine. I was wondering why it was prescribed.*” Another woman emphasized the importance of receiving this information: “*What are its benefits? What happens to us when we take it? How does it work in our body?”* Pharmacist counselling at the point of dispensing provided additional adherence support.

At the *health-system level*, access, affordability, and service organization shaped uptake. LDA was generally perceived as affordable and its reported availability supported access; however, financial constraints remained a barrier for some women, particularly those with limited resources. Requiring a provider prescription to obtain LDA through pharmacies or health posts limited ease of access by requiring formal health system contact, while also supporting clinical oversight of use. Once prescribed, consistent availability and co-located supply at points of care reduced additional barriers to obtaining the medication.

**Social opportunity** at the *patient-level* shaped acceptance and sustained uptake of LDA. Low community awareness of LDA limited normative understanding of its preventive role, while family and peer influences could either discourage or reinforce uptake through the spread of concerns (e.g., inaccurate information about safety) or supportive experiences (e.g., shared experiences with peers). Family support, including reminders, assistance obtaining medication, and financial or logistical help, facilitated use. As one woman explained, “*My family is supportive, which makes it easier for me…they do everything—from helping with household tasks to getting the medicine.”*

Trust in health information was also shaped by social hierarchies, with patients often preferring advice provided by doctors and placing greater weight on guidance from male doctors than female doctors (frequently misidentified as nurses), reflecting broader gendered perceptions of provider credibility. Despite limited awareness, LDA was generally viewed as socially acceptable, with less stigma than other pregnancy medications such as antihypertensives, facilitating uptake when women understood its purpose.

**Reflective motivation** at the *patient-level* shaped women’s perceptions of LDA’s necessity, safety, and value during pregnancy. Concerns about potential harm to the fetus, broader beliefs against medication use in pregnancy, preference for home remedies, and uncertainty about long-term effects reduced motivation to initiate or continue LDA. Additional concerns included the perceived need and burden of adding another medication alongside existing supplements, doubts about LDA’s necessity compared with familiar pregnancy supplements such as iron and calcium, and perceptions that LDA was less essential because it was not universally provided free of cost. As one woman explained:

> *“Ah, sometimes people get confused…Should I really take this medicine? I have been taking iron and calcium regularly, so now, is it necessary to take aspirin as well? That question often comes to mind”* [Pregnant woman #7].

Conversely, trust in healthcare providers and clear clinician recommendations strongly facilitated uptake by increasing the perceived legitimacy and necessity of LDA. As many participants emphasized, “*everyone trusts the doctor.*” One woman further explained:

> *“After the doctor properly explains how aspirin works, we will be convinced, and that is why we take the medicine…because of the doctor’s convincing power, we, the patients, continue taking the medicine”* [Pregnant woman #7].

Belief in LDA’s effectiveness, awareness of pre-eclampsia risks, responsibility to protect the fetus, and previous positive experiences during pregnancy with LDA further strengthened motivation to initiate and sustain use. As one woman explained, “*If it keeps my blood pressure under control, I will take it. If I were concerned about my health and the baby’s health, I would take it.*”

**Automatic motivation** at the *patient-level* was supported by existing medication-taking routines for pregnancy supplements such as iron and calcium, which provided established habits that could facilitate incorporation of LDA into daily practices.

## Discussion

This study provides novel insight into the many determinants shaping adoption and uptake of LDA for pre-eclampsia prevention in Nepal. While extensive evidence has established the effectiveness of LDA and documented persistent gaps in its implementation globally, emerging work has conceptualized LDA implementation as a multistep pathway [47]. Building on this work, our findings extend the prevention pathway upstream to ANC engagement and provide empirical insight into how determinants across interdependent stages shape progression from timely care engagement and risk identification to LDA prescribing and uptake. Although LDA is available, low-cost, and acceptable to women once prescribed, its impact is constrained when gaps in these upstream stages prevent women from progressing through the prevention pathway.

Timely ANC engagement represented the first critical bottleneck in the prevention pathway. Delayed ANC initiation and inconsistent attendance reduced opportunities for pre-eclampsia risk assessment and LDA initiation within the effective gestational window, consistent with evidence from LMIC settings demonstrating that late entry into care limits access to preventive and diagnostic interventions [23,48,49]. In Nepal, these delays reflected interacting patient- and health-system level determinants across COM-B components, including transportation barriers, costs of care, household responsibilities, gendered decision-making dynamics, and limited awareness of pregnancy risks and the importance of ANC engagement – determinants consistently reported as barriers to ANC engagement across LMIC settings [49,50]. Our findings extend this evidence by illustrating how these barriers create upstream constraints that shape progression through the pre-eclampsia prevention pathway, limiting subsequent opportunities for risk identification, prescribing, and LDA uptake.

Among women who accessed ANC, screening to identify women at risk for pre-eclampsia was a major implementation bottleneck. While routine ANC platforms provided opportunities for screening through blood pressure measurement, urine testing, and clinical assessment [51], our findings suggest that these processes were inconsistently implemented due to interacting determinants across COM-B components, particularly limited provider capability and constrained physical opportunity within health services. Variability in basic screening practices, including concerns regarding blood pressure measurement quality, reflects broader evidence from LMICs showing that blood pressure measurement is not always performed, and when undertaken it is not consistently conducted using standardized techniques [52]. Beyond routine screening, the use of more advanced approaches such as uterine artery Doppler were limited by cost, equipment availability, and access to trained personnel, consistent with FIGO guidance highlighting that although Doppler-based approaches improve predictive accuracy for first-trimester pre-eclampsia screening, their resource requirements limit uptake in LMICs [53].

Following risk identification, LDA prescribing represented a transition from clinical eligibility to preventive action. At this stage, adoption was less constrained by provider motivation and more by the extent to which the health system provided the physical opportunity to support standardized prescribing. Although providers generally recognized the value of LDA and expressed willingness to prescribe, prescribing remained dependent on individual clinical judgement and local capacity in the absence of national guidance, standardized protocols, and clear delivery pathways. These system-level constraints may be compounded by broader workforce challenges, including burnout, which can affect providers’ capacity and motivation to deliver preventive care [54]. However, our findings suggest that addressing provider motivation alone is insufficient; prevention must also be institutionalized within routine ANC systems. Evidence from implementation research in LMICs similarly demonstrates that clinical recommendations alone rarely translate into routine practice without supporting mechanisms, including training, supervision, procurement systems, and integration into routine service delivery [55].

LDA uptake among pregnant women appeared to be less constrained by medication availability or acceptability than by the conditions under which prescribing occurred. Women generally accepted LDA when its purpose and safety were clearly explained, while limited knowledge, concerns about medication use during pregnancy, and uncertainty regarding preventive benefits influenced uptake. Consistent with broader implementation literature, LDA use during pregnancy is shaped by interacting patient, provider, and health-system determinants, including communication, trust, risk perception, and access to care, rather than individual motivation alone [56]. Provider counselling emerged as a key mechanism linking prescribing decisions to sustained medication use, highlighting that uptake depends not only on whether LDA is prescribed, but on how preventive care is communicated and supported within routine ANC encounters.

### Strengths and Limitations

This study has several strengths. First, it incorporates perspectives from multiple stakeholders involved across the prevention pathway, including pregnant women, healthcare providers, and MCH stakeholders, allowing us to understand how determinants are experienced and understood by different actors. Second, the inclusion of three tertiary hospitals with differing institutional contexts enhanced contextual diversity. Third, use of the COM-B model provided a theory-informed approach to identifying and organizing determinants, while the pathway-based analysis moved beyond classification of barriers and facilitators by examining how determinants interact across sequential stages of prevention.

This study also benefited from purposive sampling across participant groups and hospital settings, alongside an iterative assessment of data sufficiency throughout data collection and analysis. Thematic saturation was considered alongside the breadth of findings across participant groups and hospital sites when determining whether the data were sufficient to address the study objectives. This approach reflects broader debate regarding the use of saturation as a marker of qualitative sufficiency and rigor, particularly in complex, multi-site health systems research where variation across actors and settings is an important feature of the phenomenon under study [42–45].

Analytic rigor was further supported by standardized templates and stepwise procedures that enabled consistency, transparency, and traceability throughout data reduction and interpretation, while enabling simultaneous engagement by a multidisciplinary team with varying qualitative expertise [42]. Involvement of bilingual Nepali team members in data collection and interpretation strengthened contextual understanding and helped identify linguistic, cultural, and contextual nuances, while researchers from different backgrounds brought additional perspectives to interpretation.

Several limitations should be considered. While the findings may be transferable to ANC in tertiary hospital settings, transferability to community settings and populations with less engagement with formal ANC services may be limited [42]. Further, perspectives of other actors who may influence LDA uptake, such as husbands and mothers-in-law, were not captured, potentially limiting our understanding of broader social influences on uptake. Additionally, all interviewed pregnant women reported literacy, potentially underrepresenting women experiencing greater informational barriers. These barriers may be even more pronounced among women with less access to information and resources; thus, our findings may underestimate the extent of these barriers in more disadvantaged populations.

Healthcare provider interviews may also have been influenced by social desirability bias, particularly given the professional context of discussions about clinical practice. This was mitigated through private interviews, assurance of confidentiality, and the use of open-ended questions and probes to encourage detailed discussion of providers’ experiences and practices. Although informal observations of clinic settings provided contextual understanding and informed interpretation of interview data, they were not systematically recorded or analyzed; future research incorporating direct observation could provide additional insight into routine care practices. Finally, implementation contexts evolve rapidly. Changes observed during data collection, for example, the introduction of new screening and prescribing protocols at one of our study hospitals, highlight the challenge of capturing implementation processes in real time and underscore the need for approaches capable of responding to rapidly evolving service delivery contexts while maintaining analytic rigor.

### Implications for research, practice, and policy

The findings highlight the need to strengthen the systems through which LDA prevention is delivered, rather than focusing solely on changing individual patient or provider behaviors. At the policy level, this includes incorporating LDA into national ANC guidelines, aligning recommendations with essential medicine and procurement systems, and establishing training and supervision mechanisms to support delivery. At the health facility level, implementation strategies should support early ANC engagement, strengthen routine risk screening, establish standardized risk assessment-to-prescribing pathways, clarify provider roles, and integrate LDA counselling into routine ANC. Together, these approaches can strengthen the interdependent stages of the pre-eclampsia prevention pathway and reduce missed opportunities for prevention.

More broadly, the findings highlight an important consideration in how preventive interventions are targeted and delivered within antenatal care. Risk-based approaches to LDA depend on timely care and accurate risk identification, whereas universal provision could reduce losses associated with missed identification but would expose more women to treatment with potentially lower expected benefit and greater cumulative risk of adverse effects [57]. Similar trade-offs have been examined for other antenatal interventions, including iron supplementation, where decisions regarding targeted versus broader delivery must consider potential benefits, risks, adherence, and health-system capacity [58].

Improving adoption and uptake will require a set of multilevel strategies that address capability, opportunity, and motivation across patients, providers, and health-system levels. The determinants identified in this study provide an empirical foundation for co-designing implementation strategies to improve LDA adoption and uptake in Nepal and potentially other settings facing similar challenges in translating LDA and other evidence-based maternal health interventions into routine practice.

In the next phases of this work, we will co-design and evaluate implementation strategies to improve LDA adoption and uptake in Nepal. Future research should build on these efforts by operationalizing and quantifying the full pre-eclampsia prevention cascade to determine how many women progress through each stage and whether upstream gaps constrain downstream LDA prescribing and uptake. These measures could then be used to evaluate whether implementation strategies improve performance at each stage and translate into gains across the cascade. Research should also examine how implementation strategies perform across different health-systems and community contexts and how they can be adapted to settings with different implementation challenges and opportunities.

## Conclusion

This study demonstrates that LDA adoption and uptake for pre-eclampsia prevention are shaped by interdependent determinants across the prevention pathway, extending from early ANC engagement and risk identification to provider prescribing and pregnant women’s uptake. While LDA is acceptable, available, and supported by strong evidence, its impact may be constrained when gaps at upstream stages prevent progression through the prevention pathway. Improving LDA adoption and uptake will require coordinated, multilevel implementation strategies that strengthen policies, service delivery systems, provider capacity, and patient communication. These findings provide context-specific evidence to inform efforts to integrate LDA into maternal health systems in Nepal and other settings seeking to translate evidence-based preventive interventions into routine care.

## Data availability

The qualitative interview transcripts generated and analyzed during the current study are not publicly available due to ethical and privacy considerations related to the potential identification of participants. De-identified data may be available from the corresponding author on reasonable request, subject to approval from the relevant ethics committees and appropriate data protection safeguards.

## Author contributions

LH: Conceptualization, Data Curation, Formal Analysis, Investigation, Methodology, Project Administration, Writing – Original Draft Preparation. ECR and AS: Conceptualization, Methodology, Supervision, Writing – Review & Editing. JD: Conceptualization, Methodology, Writing – Review & Editing. SU and AO: Investigation, Formal Analysis, Writing – Review & Editing. TR and PP: Formal Analysis, Writing – Review & Editing. All authors reviewed and edited the manuscript, approved the final version, and agreed to be accountable for all aspects of the work.

## Acknowledgements

We are deeply grateful to the women, healthcare providers, and maternal and child health stakeholders who generously shared their time, experiences, and perspectives. We recognize the women who contributed while seeking care in busy health facilities and the healthcare providers and stakeholders who participated alongside their clinical and professional responsibilities. Their contributions were essential to advancing understanding of pre-eclampsia prevention.

## Supporting information

**S1 File. Complete set of determinants across the pre-eclampsia prevention pathway**. Four supplementary tables presenting the complete set of determinants identified for each behavior in the pre-eclampsia prevention pathway, organized by COM-B component, level, and whether each determinant was identified as a barrier and/or facilitator.

## References

1. Ramchandani J, Garg J, Rajendran G, Aronow WS, Frishman WH, Gupta CA. Hypertensive Disorders of Pregnancy: A Review of the Current Literature and Future Directions. Cardiology in Review. 2025; doi: 10.1097/CRD.0000000000000870.

2. Ru Fu MM, Yihui Li MM, Xiaogang L, Weihong J. Hypertensive Disorders in Pregnancy: Global Burden From 1990 to 2019, Current Research Hotspots and Emerging Trends. Current Problems in Cardiology. 2023;48(12):101982.

3. Wang W, Xie X, Yuan T, Wang Y, Zhao F, Zhou Z, et al. Epidemiological trends of maternal hypertensive disorders of pregnancy at the global, regional, and national levels: a population-based study. BMC Pregnancy Childbirth. 2021;21(1):364.

4. Cresswell J, Alexander M, Chong MYC, Link HM, Pejchinovska M, Gazeley U, et al. Global and regional causes of maternal deaths 2009-20: a WHO systematic analysis. The Lancet Global Health. 2025;13(4):e626–e634.

5. World Health Organization (WHO). Pre-eclampsia. Geneva: World Health Organization; 2025. https://www.who.int/news-room/fact-sheets/detail/pre-eclampsia. Accessed 27 July 2026.

6. Thapa T, Sharma S, Sigdel D, Silwal K, Joshi A. Pregnancy Induced Hypertension among Pregnant Women Delivering in a Tertiary Care Hospital: A Descriptive Cross-sectional Study. JNMA J Nepal Med Assoc. 2021;59(244):1209–1214.

7. Ministry of Health and Population (MoHP), National Statistics Office (NSO). National Population and Housing Census 2021: Nepal Maternal Mortality Study 2021. Kathmandu: Ministry of Health and Population and National Statistics Office; 2022.

8. Das S, Das R, Bajracharya R, Baral G, Jabegu B, Odland J, et al. Incidence and Risk Factors of Pre-Eclampsia in the Paropakar Maternity and Women’s Hospital, Nepal: A Retrospective Study. Int J Environ Res Public Health. 2019;16(19):3571.

9. Mustary M, Ansariadi, Syam A, Riskiyani S, Erika KA, Moedjiono AI, et al. Preeclampsia: Etiology, Pathophysiology, Risk Factors, Impact and Prevention: A Narrative Review. Iran J Public Health. 2024;53(11):2392–2403.

10. Turbeville HR, Sasser JM. Preeclampsia beyond pregnancy: long-term consequences for mother and child. Am J Physiol Renal Physiol. 2020;318(6):F1315–F1326.

11. Henderson JT, Whitlock EP, O’Connor E, Senger CA, Thompson JH, Rowland MG. Low-dose aspirin for prevention of morbidity and mortality from preeclampsia: a systematic evidence review for the U.S. Preventive Services Task Force. Ann Intern Med. 2014;160(10):695–703.

12. Ruano R, Fontes RS, Zugaib M. Prevention of preeclampsia with low-dose aspirin – a systematic review and meta-analysis of the main randomized controlled trials. Clinics. 2005;60(5):407–414.

13. Wang Y, Guo X, Obore N, Ding H, Wu C, Yu H. Aspirin for the prevention of preeclampsia: A systematic review and meta-analysis of randomized controlled studies. Front Cardiovasc Med. 2022;9:936560.

14. Duley L, Meher S, Hunter KE, Seidler AL, Askie LM. Antiplatelet agents for preventing pre-eclampsia and its complications. Cochrane Database of Systematic Reviews. 2019(10):CD004659.

15. Sinkey RG, Battarbee AN, Bello NA, Ives CW, Oparil S, Tita ATN. Prevention, Diagnosis, and Management of Hypertensive Disorders of Pregnancy: a Comparison of International Guidelines. Curr Hypertens Rep. 2020;22(9):66.

16. Sohail H. Prophylaxis of Preeclampsia With Low-Dose Aspirin: A Comprehensive Review. Cureus. 2025;17(11):e96529.

17. Werner EF, Hauspurg AK, Rouse DJ. A Cost-Benefit Analysis of Low-Dose Aspirin Prophylaxis for the Prevention of Preeclampsia in the United States. Obstet Gynecol. 2015;126(6):1242–1250.

18. Mallampati D, Grobman W, Rouse DJ, Werner EF. Strategies for Prescribing Aspirin to Prevent Preeclampsia: A Cost-Effectiveness Analysis. Obstet Gynecol. 2019;134(3):537–544.

19. Ortved D, Hawkins TL, Johnson JA, Hyett J, Metcalfe A. Cost-effectiveness of first-trimester screening with early preventative use of aspirin in women at high risk of early-onset pre-eclampsia. Ultrasound Obstet Gynecol. 2019;53(2):239–244.

20. World Health Organization (WHO). WHO recommendations on antiplatelet agents for the prevention of pre-eclampsia. Geneva: World Health Organization; 2021. Licence: CC BY-NC-SA 3.0 IGO.

21. Foster AB, Park F, Hyett J. Do first-trimester screening algorithms for preeclampsia aligned to use of preventative therapies reduce the prevalence of pre-term preeclampsia: A systematic review and meta-analysis. Prenat Diagn. 2023;43(7):950–958.

22. Baschat AA, Magder LS, Doyle LE, Atlas RO, Jenkins CB, Blitzer MG. Prediction of preeclampsia utilizing the first trimester screening examination. Am J Obstet Gynecol. 2014;211(5):514.E1–514.E7.

23. World Health Organization (WHO). WHO recommendations on antenatal care for a positive pregnancy experience. Geneva: World Health Organization; 2016. Licence: CC BY-NC-SA 3.0 IGO.

24. Cross-Barnet C, Courtot B, Benatar S, Hill I. Preeclampsia Risk and Prevention among Pregnant Medicaid Beneficiaries. J Health Care Poor Underserved. 2020;31(4):1634–1647.

25. Lake ES, Ayele M, Tilahun BD, Erega BB, Belay AS, Yilak G. Obstetric care provider’s knowledge about the use of low dose aspirin for preeclampsia prevention in low and middle income countries: a systematic review and meta-analysis. BMC Pregnancy Childbirth. 2024;24(1):611.

26. Vinogradov R, Smith V, Hiu S, McParlin C, Farnworth A, Araújo-Soares V. Let’s talk aspirin: A survey of barriers and facilitators faced by midwives when engaging in conversations about aspirin with women at risk of pre-eclampsia. Midwifery. 2023;127:103860.

27. Sium AF, Gudu W, Tolu LB, Birara M, Bekele D. Missed opportunity for aspirin prophylaxis for preeclampsia prevention: a cross-sectional study from Sub-Saharan Africa. AJOG Glob Rep. 2023;4(1):100295.

28. van Montfort P, Scheepers HCJ, van Dooren IMA, Meertens LJE, Zelis M, Zwaan IM, et al. Low-dose aspirin usage among women with an increased preeclampsia risk: A prospective cohort study. AOGS. 2020;99(7):811–955.

29. Gittens S, Pak J. Use of Daily Low-Dose Aspirin in Pregnancy for Women at High Risk of Preeclampsia at Georgetown Public Hospital Corporation. International Journal of Research and Review. 2022;9(9):339–348.

30. Combs CA, Kumar NR, Morgan JL, and SMFM Patient Safety and Quality Committee. Society for Maternal-Fetal Medicine Special Statement: Prophylactic low-dose aspirin for preeclampsia prevention quality metric and opportunities for quality improvement. American Journal of Obstetrics & Gynecology. 2023;229(2):B2–B9.

31. Olson DN, Russell T, Ranzini AC. Assessment of Adherence to Aspirin for Preeclampsia Prophylaxis and Reasons for Nonadherence. Am J Obstet Gynecol. 2022;4(5):100663.

32. Lorreia ADA, Renaud S, Pimentel VM. Aspirin Acceptance Rate and Perceived Preeclampsia Risk after the Implementation of Universal Screening for Preeclampsia. American Journal of Obstetrics and Gynecology. 2021;224(2):S184.

33. World Health Organization (WHO). Web Annex A. World Health Organization Model List of Essential Medicines – 23rd List, 2023. In: The selection and use of essential medicines 2023: Executive summary of the report of the 24th WHO Expert Committee on the Selection and Use of Essential Medicines, 24 – 28 April 2023. Geneva: World Health Organization; 2023. (WHO/MHP/HPS/EML/2023.02). Licence: CC BYNC-SA 3.0 IGO.

34. Khanal S, Veerman L, Ewen M, Nissen L, Hollingworth S. Availability, Price, and Affordability of Essential Medicines to Manage Noncommunicable Diseases: A National Survey From Nepal. Inquiry. 2019;56:46958019887572.

35. Government of Nepal, Ministry of Health and Population, Family Welfare Division. Nepal Safe Motherhood and Newborn Health Road Map 2030. Kathmandu, Nepal; 2019.

36. Thapa K, Sanghvi H, Rawlins B, Karki YB, Regmi K, Aryal S, et al. Coverage, compliance, acceptability and feasibility of a program to prevent pre-eclampsia and eclampsia through calcium supplementation for pregnant women: an operations research study in one district of Nepal. BMC Pregnancy Childbirth. 2016;16(1):241.

37. Michie S, Atkins L, West R. The Behaviour Change Wheel: A Guide to Designing Interventions. London: Silverback Publishing; 2014.

38. Braun V, Clarke V. Thematic analysis: A practical guide. London: Sage Publications; 2022.

39. Hennink M, Hutter I, Bailey A. Qualitative research methods 2nd ed. Thousand Oaks, California USA: Sage Publications; 2020.

40. Tong A, Sainsbury P, Craig J. Consolidated criteria for reporting qualitative research (COREQ): a 32-item checklist for interviews and focus groups. International Journal for Quality in Health Care. 2007;19(6):349–357.

41. Hennink MM, Kaiser BN, Marconi VC. Code saturation versus meaning saturation: how many interviews are enough? Qual Health Res. 2017;27(4):591–608.

42. Nevedal AL, Kowalski CP, Finley EP, Fix GM, Hamilton AB, Koenig CJ. Optimizing qualitative methods in implementation research: a resource for editors, reviewers, authors, and researchers to dispel ten common misperceptions about qualitative research methods. Implementation Sci. 2026;21(4).

43. Roberts SJ, Saylor MA, Ivynian S, DiGiacomo M. Data saturation and information power: sample size in qualitative research—when is enough and how to assess? European Journal of Cardiovascular Nursing. 2026;25(5):1069–1074.

44. Mikhaeil CA, Robey D. When is enough enough? A critical assessment of data adequacy in IS qualitative research. Information and Organization. 2024;34(4):100540.

45. Sims D, Cilliers F. Assessing Data Adequacy in Qualitative Research Studies. Academic Medicine. 2025;100(6):758.

46. Kowalski CP, Nevedal AL, Finley EP, Young JP, Lewinski AA, Midboe AM, et al. Planning for and Assessing Rigor in Rapid Qualitative Analysis (PARRQA): a consensus-based framework for designing, conducting, and reporting. Implementation Sci. 2024;19(71).

47. Teal EN, Gyamfi-Bannerman C. Disparities in low dose aspirin use for preeclampsia risk reduction and opportunities for improvement. Semin Perinatol. 2026;152255.

48. Gage A, Conrad M, Knight M, Banke-Thomas A, Bintz C, Bienhoff K, et al. Measuring antenatal care timing and content across 131 low-income and middle-income countries, 1995–2023: a systematic analysis of trends. The Lancet Global Health. 2026;14,e702–e713.

49. Ali ST, Rizvi SA, Talat M, Abuzar S, Azhar M, Rehman M. Barriers to timely and adequate antenatal care: a systematic review of socioeconomic, cultural, psychosocial, and health-system factors across high and low resource settings. BMC Pregnancy Childbirth. 2026;26(96).

50. Markos Z, Kassegn A, Tesfaye D, Samuel A, Dinkneh R. Delayed initiation of antenatal care and its determinants in Ethiopia: A systematic review and meta-analysis. Midwifery. 2026;156,104734.

51. Government of Nepal, Ministry of Health and Population, Department of Health Services, Family Welfare Division. National Medical Standard for Maternal and Newborn Care: Volume III. Kathmandu, Nepal; 2022.

52. Rawlins B, Plotkin M, Rakotovao JP, Getachew A, Vaz M, Ricca J, et al. Screening and management of pre-eclampsia and eclampsia in antenatal and labor and delivery services: findings from cross-sectional observation studies in six sub-Saharan African countries. BMC Pregnancy Childbirth. 2018;18(346).

53. Poon LC, Shennan A, Hyett JA, Kapur A, Hadar E, Divakar H, et al. The International Federation of Gynecology and Obstetrics (FIGO) initiative on pre-eclampsia: A pragmatic guide for first-trimester screening and prevention. Int J Gynecol Obstet. 2019;145(Suppl 1):1–33.

54. Hazfiarini A, Corona MV, Ninckas N, Homer CSE, Turner T, Islamiah R, et al. Well-being of health workers providing maternal and newborn care: A qualitative evidence synthesis. PLOS Glob Public Health. 2026;6(2):e0005522.

55. Breneol S, Curran JA, Marten R, Minocha K, Johnson C, Wong H, et al. Strategies to adapt and implement health system guidelines and recommendations: a scoping review. Health Res Policy Sys. 2022;20(64).

56. Lee YJ, Kowaleswki L, Gould J, du Plessis L, Oshiro B, Moua M, et al. Systemic Health Intervention Design through Participatory Modeling: Low-dose Aspirin for Women at Risk of Preeclampsia. Fortune Journal of Health Sciences. 2025;8(3):798–816.

57. Wright D, Wright A, Tan MY, Nicolaides KH. When to give aspirin to prevent preeclampsia: application of Bayesian decision theory: application of Bayesian decision theory. American journal of obstetrics & gynecology. 2022;226(2):S1120–S1125.

58. Watt A, Eaton H, Eastwick-Jones K, Thomas ET, Pluddemann A. The benefits and harms of oral iron supplementation in non-anaemic pregnant women: a systematic review and meta-analysis. Fam Pract. 2025;42(1):cmae079.

